# When the storm is the strongest: Healthcare staff’s health conditions and job satisfaction and their associated predictors during the epidemic peak of COVID-19

**DOI:** 10.1101/2020.04.27.20082149

**Authors:** Stephen X. Zhang, Jing Liu, Asghar Afshar Jahanshahi, Khaled Nawaser, Jizhen Li, Hadiseh Alimoradi

**Author notes:** **Corresponding author:** Stephen X. Zhang; Address: 9–28 Nexus10 Tower, 10 Pulteney St, Adelaide SA 5000, Australia. **CRediT Author Statement:** **S. X. Zhang.:** Conceptualization, Investigation, Methodology, Formal analysis, Visualization, Writing – Original, Writing - Review & Editing, Supervision **J. Liu:** Visualization, Writing - Original Draft, Writing - Review & Editing **A.A. Jahanshahi:** Investigation (data collection), Resources, Conceptualization, Writing - Review & Editing **K. Nawaser**: Investigation (data collection) **J. Li**: Writing - Review & Editing; Funding acquisition **H. Alimoradi**: Investigation (data collection).

## Abstract

This study reports the physical health, mental health, anxiety, depression, distress, and job satisfaction of healthcare staff in Iran when the country faced its highest number of total active COVID-19 cases. In a sample of 304 healthcare staff (doctors, nurses, radiologists, technicians, etc.), we found a sizable portion reached the cutoff levels of disorders in anxiety (28.0%), depression (30.6%), and distress (20.1%). Age, gender, education, access to PPE (personal protective equipment), healthcare institutions (public vs. private), and individual status of COVID-19 infection each predicted some but not all the outcome variables of SF-12, PHQ-4, K6, and job satisfaction. The healthcare workers varied greatly in their access to PPE and in their status of COVID-19 infection: negative (69.7%), unsure (28.0%), and positive (2.3%). The predictors were also different from those identified in previous studies of healthcare staff during the COVID-19 crisis in China. This study helps to identify the healthcare staff in need to enable more targeted help as healthcare staff in many countries are facing peaks in their COVID-19 cases.

## 1. Introduction

The first COVID-19 confirmed case in Iran appeared on 19 February 2020, and four days later, a 25-year-old nurse died of COVID-19, even though her positive infection test result arrived a week after her death. Iran quickly became one of the most affected countries by total COVID-19 cases and fatalities, overwhelming the country’s healthcare staff. This study aims to report the health conditions (SF-12, K6, PHQ-4) and job satisfaction of healthcare staff during the height of the COVID-19 pandemic when Iran reached its highest count of total active cases. We also identify the risk factors to screen for healthcare staff in greater need to prioritize mental health services.

Two early studies focused on only the frontline medics dealing with COVID-19 patients directly (Xiao et al., 2020; Zhu et al., 2020). Recently, Spoorthy (2020) reviewed the early evidence on the mental health conditions of healthcare staff. The review identified five quantitative papers on the topic, and four of them examined healthcare staff in Hubei province, the initial epicenter of the outbreak in China (Cai et al., 2020; Kang et al., 2020; Lai et al., 2020; Xiao et al., 2020), and one study conducted a cross-sectional survey of 59 doctors and nurses in Guangdong province – the second most affected province in China (Liang et al., 2020). Despite such pioneering research, healthcare staff in Hubei faced a unique situation, as the coronavirus was initially unknown, and even the vast majority of the medical staff did not know what they were facing, calling it “Wuhan pneumonia”. However, medical staff in other countries were well aware of COVID-19 when it later reached them. Moreover, countries vary greatly in their healthcare systems, and we still lack quantitative evidence on the conditions of healthcare staff working in a major COVD-19 outbreak area outside of China (Spoorthy, 2020). Lastly, while this unprecedented crisis heightens everyone’s appreciation of the profession of healthcare workers, no study to date has examined their job satisfaction during the outbreak, even though job satisfaction is a critical motivational resource for healthcare workers to prevent burnout (Zhang et al., 2020a) and get through the pandemic.

This study reports not only the health conditions but also the job satisfaction of healthcare staff in Iran during a critical moment when Iran reached its peak in total active COVID-19 cases. This study helps to enable evidence-based identification of healthcare staff in need of more targeted psychiatric assistance via telephone, internet, and mobile apps (Kang et al., 2020) at a time when more and more countries are reaching their COVID-19 epidemic peaks.

## 2. Methods

### 2.1 Settings

The data collection took place on April 5–20, 2020 during the COVID-19 pandemic in Iran. On April 5, the date we started our survey of healthcare staff, the number of total active COVID-19 cases in Iran peaked at 32,612 cases, there were 2,483 daily new cases, and 3,603 total fatalities, 43 of whom were healthcare workers (Gharebaghi & Heidary, 2020).

News reported that many healthcare workers in Iran did not have hazmat suits and were short of latex and sterile gloves (Alijani, 2020). A doctor described the situation facing her and her colleagues: “They just give us latex gloves – two pairs per shift. Even if we want to protect ourselves, we have to buy our own protective suits. If we manage to track some down, the prices are crazy! I bought nine suits for 2.5 million toman [150 euro]. … Some of our colleagues, especially nurses, have stopped coming to work. We can’t go on like this for long. The psychological pressure is unbearable. Every second you think you might get infected yourself and die”(Alijani, 2020).

Such a situation was corroborated by our primary interviews. We conducted short interviews by telephone with five doctors and nurses in five different public and private healthcare facilities in Iran during the study period. The interviews revealed that healthcare workers varied in their access to PPE. Due to the huge number of patients who rushed in suspecting they had COVID-19 and the high number of infected cases, the healthcare workers had very long working hours (some reported working 19 hours in a row a day), and most of them had been away from their family for over a month out of concern about bringing the virus home.

The participants of our survey included healthcare staff, such as doctors, nurses, radiology technologists, obstetrics and healthcare administrators in public and private hospitals in Iran. The study was approved by Tsinghua University (#20200322), and a total of 304 healthcare staff participated in this data collection.

### 2.2 Measures

The survey asked healthcare staff demographic information on their age, gender, marital status, education level, and job function, as well as physical health, mental health, anxiety, depression, distress, job satisfaction, access to PPE, whether they worked in a private or public healthcare organization, and their status of COVID-19 infection.

*Physical health and mental health*. We used SF-12, a widely applied scale to evaluate individuals’ physical health and mental health status (Ware Jr et al., 1996). SF-12 contains eight subscales, which are calculated into a physical health composite score (PCS) and a mental health composite score (MCS).

*Depression and anxiety*. Depression and anxiety were assessed by PHQ-4 (Patient Health Questionnaire-4), which is often used to assess depression and anxiety for the general public (Löwe et al., 2010). PHQ-4 contains a 2-item scale of depression (Kroenke et al., 2003) and a 2-item scale of anxiety (Kroenke et al., 2007).

*Distress*. We assessed distress by the 6-item Kessler psychological distress scale (K6) (Kessler et al., 2002). K6 is a self-administrated scale to screen for mental health problems among the general public. The Cronbach’s alpha was 0.91.

*Job satisfaction*. We used the Brayfield–Rothe job satisfaction scale (Brayfield & Rothe, 1951; Montazeri et al., 2011; Montazeri et al., 2009; Rohani et al., 2010), which contains five items: “I feel fairly satisfied with my present job”; “ Most days I am enthusiastic about my work”; “Each day at work seems like it will never end”; “I find real enjoyment in my work”; “I consider my job to be rather unpleasant.” For each item, the participants answered to what extent they agreed with the statement. All responses were made on a 5-point Likert scale (1= *totally disagree* to 5= *totally agree*). The Cronbach’s alpha was 0.83.

### 2.3 Statistical analysis

All the analyses were done in Stata 16.0 at a significance level of 0.05. The categorical variables were tabulated by the frequency and percentage, and continuous variables were summarized by mean ± standard deviation in the descriptive statistics (Table 1). To identify the potential risk factors for health conditions (SF-12, K6, PHQ-4) and job satisfaction of the healthcare staff, we used linear regression analysis and interpreted the coefficient, 95% CI, and p-value.

**Table 1.** Descriptive statistics for healthcare staff in Iran during the height of the COVID-19 epidemic (N=304)

| Variables |  | mean + SD or No. (%) |
| --- | --- | --- |
| Age (years) |  | 35.1±9.1 |
| Gender | Male | 126 (41.4%) |
|  | Female | 178 (58.6%) |
| Marital status | Single | 114 (37.5%) |
|  | Married without child | 57 (18.7%) |
|  | Married with one child | 61 (20.0%) |
|  | Married with more than one child | 69 (22.7%) |
|  | Divorced | 3 (1.0%) |
| Highest educational level | Under diploma | 7 (2.3%) |
|  | Diploma (12 years) | 21 (6.9%) |
|  | 2-years college | 37 (12.2%) |
|  | Graduated or studying a bachelor degree | 143 (47.0%) |
|  | Graduated or studying a master degree | 43 (14.1%) |
|  | Graduated or studying a doctoral degree | 53 (17.4%) |
| Working in a public or private institution | Public | 223 (73.4%) |
|  | Private | 81 (26.6%) |
| Job function | Radiology technologists | 64 (21.1%) |
|  | Nurses | 63 (20.7%) |
|  | Doctors | 45 (14.8%) |
|  | Healthcare administrators | 39 (12.8%) |
|  | Obstetrics | 33 (10.9%) |
|  | Technicians | 30 (9.9%) |
|  | Medical students/interns | 12 (3.9%) |
|  | Supporting staff (e.g. facility/cleaning staff) | 10 (3.3%) |
|  | Volunteers | 8 (2.6%) |
| Access to PPE (Personal Protective Equipment) | Never | 14 (4.8%) |
|  | Rarely | 45 (15.4%) |
|  | Sometimes | 89 (30.5%) |
|  | Very often | 78 (26.7%) |
|  | Always | 66 (22.6%) |
| COVID-19 infection status | Negative | 212 (69.7%) |
|  | Unsure | 85 (28.0%) |
|  | Positive | 7 (2.3%) |
| Physical health score (PCS of SF-12) |  | 40.7 ± 7.0 |
| Mental health score (MCS of SF-12) |  | 26.3 ± 7.5 |
| Anxiety score (PHQ-4) |  | 2.0 ± 1.5 |
| Depression score (PHQ-4) |  | 2.1 ± 1.4 |
| Distress score (K6: 1–5 anchor) |  | 14.8 ± 5.3 |
| Job satisfaction |  | 3.3 ± 0.7 |

## 3. Results

### 3.1 Descriptive findings

Table 1 presents the descriptive findings. Of the 304 healthcare staff surveyed, 58.6% (178) were female, and 41.4% (126) were male. Over half of them, 55.9% (170), were aged in their 30s or 40s. The majority (73.4%) of the healthcare staff worked in public healthcare facilities, and the rest 26.6% worked in private institutions. Over a quarter (22.6%) of the healthcare staff indicated they “*always*” had access to PPE. The rest indicated their access to PPE was “*very often*” (26.7%), “*sometimes*” (30.5%), or “*rarely*” (15.4%). The remaining 4.8% of the healthcare staff unfortunately “*never*” had access to PPE. Of all the healthcare staff, 69.7% (212) reported they were negative for COVID-19 infection, and 2.3% (7) of them had reported positive. The remaining 28.0% (85) were unsure, likely due to the shortage of testing.

The healthcare staff averaged 40.7 ± 7.0 (st.d.) on the physical health composite score (PCS of SF-12) and 26.3 ± 7.5 on the mental health composite score (MCS of SF-12). The mean of anxiety (GAD-2) score was 2.0 ± 1.5 and 28.0% (85) of the healthcare staff reached the cutoff score of 3, a level that should prompt “a clinical interview to determine whether a mental disorder is present” (Spitzer, Williams, & Kroenke, 1990). The mean score of depression (PHQ-2) was 2.1 ± 1.4 and 30.6% (93) reached the cut-off score of 3, indicating they would need a clinical interview to determine whether depression is present. The mean K6 score was 14.8 ± 5.3 with a 1–5 scoring, and 20.1% (61) of the healthcare staff exceeded the cut-off score indicating the likely presence of a clinical distress disorder.

We tried to compare the outcome variables in our sample with published studies of the same variables among the general population in Iran during the COVID-19 crisis or among healthcare staff in Iran before the COVID-19 crisis, however we could not find such prior studies in Iran. However, there are prior validation studies of such variables among the general population in Iran for comparison. The mental health composite (MCS of SF-12) in our sample of 26.3 (7.5) was significantly lower (p<0.001) than mental health scores reported in three previous studies of 46.3 (10.4), 44.2 (10.8), and 44.6 (11.9) respectively (Montazeri et al., 2011; Montazeri et al., 2009; Rohani et al., 2010). The physical health composite (PCS of SF-12) in our sample of 40.7 (7.0) was also significantly lower (p<0.001; p<0.001; p<0.05) respectively than scores reported in the same three studies of 50.1 (8.5), 48.2 (8.2), and 42.3 (11.4).

### 3.2 The risk factors for healthcare staff’s physical health, mental health, depression, anxiety, distress and job satisfaction

Table 2 presents the regression results on the potential risk factors for healthcare staff’s physical health, mental health, anxiety, depression, distress and job satisfaction. The regression results on SF-12 show that older healthcare staff enjoyed better mental health (β=0.230, 95% CI: 0.106 to 0.355, p=0.000) but not physical health (β=-0.061, 95% CI: −0.174 to 0.052, p=0.290). Education level predicted physical health (β=0.977, 95% CI: 0.204 to 1.750, p=0.014) and mental health (β=-0.902, 95% CI: −1.757 to −0.047, p=0.039). Female staff members experienced more distress (β=2.140, 95% CI: 0.851 to 3.429, p=0.001) and depression (β=0.457, 95% CI: 0.117 to 0.796, p=0.009). Marital status failed to predict any outcome variables.

**Table 2.** Risk factors of the health conditions (SF-12, PHQ-4 and K6) and job satisfaction among healthcare staff (N=292)

| Variables |  | Physical health (SF-12) |  | Mental health (SF-12) |  | Anxiety (PHQ-4) |  | Depression (PHQ-4) |  | Distress (K6) |  | Job satisfaction |  |
| --- | --- | --- | --- | --- | --- | --- | --- | --- | --- | --- | --- | --- | --- |
| | | $\beta$ (95%CI) | p | $\beta$ (95%CI) | p | $\beta$ (95%CI) | p | $\beta$ (95%CI) | p | $\beta$ (95%CI) | p | $\beta$ (95%CI) | p |
| Age |  | -0.061 | 0.290 | 0.230 | 0.000 | -0.021 | 0.059 | -0.020 | 0.054 | -0.038 | 0.339 | -0.004 | 0.512 |
|  |  | (-0.174 to -0.052) |  | (0.106 to 0.355) |  | (-0.044 to 0.001) |  | (-0.040 to 0.000) |  | (-0.115 to 0.040) |  | (-0.015 to 0.007) |  |
| Gender |  | 1.040 | 0.283 | 0.110 | 0.918 | 0.354 | 0.060 | 0.457 | 0.009 | 2.140 | 0.001 | -0.052 | 0.574 |
|  |  | (-0.865 to 2.944) |  | (-1.995 to 2.214) |  | (-0.016 to 0.723) |  | (0.117 to 0.796) |  | (0.851 to 3.429) |  | (-0.235 to 0.130) |  |
| Marital status |  | 0.030 | 0.948 | -0.225 | 0.654 | -0.004 | 0.965 | 0.030 | 0.704 | -0.124 | 0.675 | 0.031 | 0.460 |
|  |  | (0.865 to 0.925) |  | (-1.214 to 0.764) |  | (-0.170 to 0.163) |  | (-0.124 to 0.183) |  | (-0.706 to 0.458) |  | (-0.051 to 0.113) |  |
| Education level |  | 0.977 | 0.014 | -0.902 | 0.039 | 0.058 | 0.445 | 0.115 | 0.100 | 0.084 | 0.751 | 0.026 | 0.483 |
|  |  | (0.204 to 1.750) |  | (-1.757 to -0.047) |  | (-0.091 to 0.207) |  | (-0.022 to 0.252) |  | (-0.436 to 0.604) |  | (-0.047 to 0.100) |  |
| Access to PPE<br>(Personal Protective<br>Equipment) |  | 1.875 | 0.000 | -0.404 | 0.396 | -0.069 | 0.378 | -0.084 | 0.241 | -0.570 | 0.038 | 0.150 | 0.000 |
|  |  | (1.027 to 2.724) |  | (-1.342 to 0.534) |  | (-0.223 to 0.085) |  | (-0.226 to 0.057) |  | (-1.107 to -0.033) |  | (0.074 to 0.226) |  |
| Public or<br>private<br>institution | Public | reference |  |  |  |  |  |  |  |  |  |  |  |
|  | Private | 0.193 | 0.856 | 2.349 | 0.046 | -0.128 | 0.520 | -0.006 | 0.973 | 0.603 | 0.387 | -0.079 | 0.426 |
|  |  | (-1.895 to 2.281) |  | (0.041 to 4.657) |  | (-0.521 to 0.264) |  | (-0.367 to 0.355) |  | (-0.768 to 1.973) |  | (-0.273 to 0.116) |  |
| COVID-19<br>infection | Negative | reference |  |  |  |  |  |  |  |  |  |  |  |
|  | Positive | -3.482 | 0.293 | 1.807 | 0.621 | 0.157 | 0.781 | -0.193 | 0.710 | 1.481 | 0.454 | 0.191 | 0.496 |
|  |  | (-9.992 to 3.027) |  | (-5.388 to 9.002) |  | (-0.956 to 1.271) |  | (-1.217 to 0.830) |  | (-2.406 to 5.367) |  | (-0.360 to 0.741) |  |
|  | Unsure | -1.081 | 0.286 | -0.727 | 0.516 | 0.688 | 0.001 | 0.449 | 0.013 | 1.649 | 0.016 | 0.260 | 0.008 |
|  |  | (-3.074 to 0.912) |  | (-2.923 to 1.476) |  | (0.303 to 1.073) |  | (0.094 to 0.803) |  | (0.304 to 2.995) |  | (-0.451 to -0.070) |  |

A healthcare worker’s status of COVID-19 infection (negative, unsure, positive) significantly predicted the outcome variables. Compared to the 69.7% of staff who indicated negative to COVID-19 infection, the 28.0% who were unsure whether they had COVID-19 infection had higher depression (β=0.449, 95% CI: 0.094 to 0.803, p=0.013), higher anxiety (β=0.688, 95% CI: 0.303 to 1.073, p=0.001), more distress (β =1.649, 95% CI: 0.304 to 2.995, p=0.016), and lower job satisfaction (β=-0.260, 95% CI: −0.451 to −0.070, p=0.008). There were no statistically significant differences between those who reported COVID-19 negative vs. positive status, likely due to the small number of individuals who were positive (7).

Institutionally, those who worked in private healthcare institutions had better mental health (β=2.349, 95% CI: 0.041 to 4.657, p=0.046). Healthcare staff’s access to personal protective equipment predicted better physical health (β=1.875, 95% CI: 1.027 to 2.724, p=0.000), lower distress (β=-0.570, 95% CI: −1.107 to −0.033, p=0.038) and better job satisfaction (β=0.150, 95% CI: 0.074 to 0.226, p=0.000).

## 4. Discussion

COVID-19 hit Iran early and hard, putting enormous pressure on the country’s healthcare staff. This study presents the first attempt to document healthcare staff’s health conditions and job satisfaction as well as their predictors during the COVID-19 pandemic. Overall, a sizable percentage of healthcare staff reached the cutoff values for mental disorder concerns on common psychiatric measures (20.1% by K6; 20.6% by PHQ-2; 28.0% by GAD-2) to require medical attention themselves. However, the predictors of these outcome variables varied, revealing more nuanced patterns.

### 4.1 The risk factors (predictors) vary across settings

The predictors in our sample from Iran differ from those in healthcare samples in earlier studies in China. Gender predicted both anxiety and depression among healthcare staff in China (Lai et al., 2020), but in our Iranian sample, gender predicted depression and distress, but not anxiety. Age also predicted anxiety and depression of healthcare staff in China (Lai et al., 2020) but not in our sample in Iran. Instead, age predicted the mental health score by SF-12 in our sample. The education level of healthcare staff in China also predicted less depression (Liu et al., 2020), a relationship that was not significant in our sample from Iran.

Taken together, the results suggest distinct patterns of the predictors of the mental health conditions of healthcare staff in Iran and China during the COVID-19 crisis. Such distinct patterns of predictors among healthcare staff across countries resonant with an earlier study of the mental health of the public in Iran (Jahanshahi et al., 2020). Jahanshahi et al. (2020) suggest the predictors differed because “different countries vary in their medical systems, the availability of personal protective equipment (PPE), cultures, labor and employment conditions, the policies of lockdown, the ease of working from home and maintaining a living in a pandemic, and the information in both mainstream and social media”. Our results, hence, corroborate their suggestions, and we suggest future research to identify useful predictors of mental health in individual countries during the COVID-19 pandemic for not only the public but also healthcare staff.

### 4.2 Unique predictors uncovered in our study

Our research also uncovered several unique risk factors. First, healthcare staff’s access to PPE was associated with better physical health and job satisfaction and less distress, suggesting PPE played a role in not only their physical health but also psychological distress and job satisfaction. These findings demonstrate the role of PPE beyond protecting healthcare staff merely physically. The findings support the argument of Cai et al. (2020) that adequate PPE motivates healthcare workers during outbreaks of infectious diseases.

Second, our findings revealed that those healthcare staff who were unsure whether they had COVID-19 were significantly more distressed, depressed, and anxious, and had lower job satisfaction. This finding implies the need to either prioritize the testing of healthcare staff if testing kits are available or otherwise to prioritize them for mental health support. It could be very harmful for healthcare staff to work in conditions of distress, depression, anxiety, and lower job satisfaction for a prolonged period in this unprecedented healthcare crisis (Zhang et al., 2020b).

Third, our results show that healthcare staff working in private institutions had better mental health (MCS of SF-12) than staff in public institutions. This finding suggests research should examine how private institutions function in a crisis compared to public institutions to identify possible areas of improvement.

### 4.3 No single universal risk factor predicted outcome variables

While risk factors in this study did predict various outcome variables from physical health to job satisfaction, we did not find a universal risk factor that predicted all outcome variables across the board. Hence, we suggest more research is needed to identify unique risk factors for the variables of concern. In doing so, we also highlight a challenge in screening mentally vulnerable populations during the ongoing COVID-19 pandemic where we may need to screen for specific mental disorders by distinct sets of risk factors. To date, psychological assistance programs have yet to make such distinctions. For instance, early mental health programs in Hubei to help the mentally vulnerable consisted of four teams: the psychosocial response team, technical support, psychiatric medics, and psychological assistance hotline teams (Kang et al., 2020), without distinguishing specific mental disorders and their associated risk factors. Psychiatrists specializing in anxiety disorder, or depression issues, for instance, would need to identify the unique risk factors for predicting their areas of concern.

### 4.4 Limitations

We aim to capture the health conditions of healthcare staff during the peak medical demand of the COVID-19 pandemic. We were fortunate that the number of total active COVID-19 cases in Iran reached its peak on the week we started the survey, and it is reasonable to think the healthcare demand during our survey period was very heavy. Still we were not able to obtain the number of COVID-19 patients in healthcare facilities in Iran. The exact number of patients, along with a longitudinal design, could reveal the fluctuations of the conditions of healthcare staff as the peak came and passed.

### 4.5 Conclusions

Protecting healthcare staff, including their physical health, mental health, and their job satisfaction for their important vocation, is paramount during the unprecedented COVID-19 pandemic. We found the risk factors for healthcare staff in Iran differed from those in past studies in China. As countries vary in their medical systems, readiness for viral epidemics, clinical capacity, and response, we encourage future studies to examine the conditions of healthcare workers and their predictors in individual countries.

## Data Availability

Upon request

## Acknowledgement

We acknowledge the support of Tsinghua University-INDITEX Sustainable Development Fund (Project No. TISD201904).

## Reference

Alijani, E. (2020, March 23). “They only give us protective gear when TV cameras come”: Iran’s doctors say they’re at risk. The Observers. https://observers.france24.com/en/20200323-Iran-Corona-protective-gear-tv-doctors

Brayfield, A. H., & Rothe, H. F. (1951). An index of job satisfaction. Journal of Applied Psychology, 35, 307–311. http://dx.doi.org/10.1037/h0055617

Cai, H., Tu, B., Ma, J., Chen, L., Fu, L., Jiang, Y., & Zhuang, Q. (2020). Psychological impacts and coping strategies of front-line medical staff during COVID-19 outbreak in Hunan, China. Medical Science Monitor, 26, e924171. http://dx.doi.org/10.12659/MSM.924171

Gharebaghi, R., & Heidary, F. (2020). COVID-19 and Iran: swimming with hands tied! Swiss Medical Weekly, 150, w20242. doi:10.4414/smw.2020.20242

Jahanshahi, A. A., Dinani, M. M., Madavani, A. N., Li, J., & Zhang, S. X. (2020). The distress of Iranian adults during the Covid-19 pandemic - More distressed than the Chinese and with different predictors. Brain, Behavior and Immunity, in production.

Kang, L., Li, Y., Hu, S., Chen, M., Yang, C., Yang, B. X., … Liu, Z. (2020). The mental health of medical workers in Wuhan, China dealing with the 2019 novel coronavirus. The Lancet Psychiatry, 7, e14. https://doi.org/10.1016/S2215-0366(20)30047-X

Kessler, R. C., Andrews, G., Colpe, L. J., Hiripi, E., Mroczek, D. K., Normand, S. L. T., … Zaslavsky, A. M. (2002). Short screening scales to monitor population prevalences and trends in non-specific psychological distress. Psychological Medicine, 32, 959–976. https://doi.org/10.1017/S0033291702006074

Kroenke, K., Spitzer, R. L., & Williams, J. B. W. (2003). The Patient Health Questionnaire-2: validity of a two-item depression screener. Medical Care, 41, 1284–1292. https://doi.org/10.1097/01.MLR.0000093487.78664.3C

Kroenke, K., Spitzer, R. L., Williams, J. B. W., Monahan, P. O., & Löwe, B. (2007). Anxiety disorders in primary care: Prevalence, impairment, comorbidity, and detection. Annals of Internal Medicine, 146, 317–326. https://doi.org/10.7326/0003-4819-146-5-200703060-00004

Lai, J., Ma, S., Wang, Y., Cai, Z., Hu, J., Wei, N., … Hu, S. (2020). Factors associated with mental health outcomes among health care workers exposed to coronavirus disease 2019. JAMA Network Open, 3, e203976. http://dx.doi.org/10.1001/jamanetworkopen.2020.3976

Liang, Y., Chen, M., Zheng, X., & Liu, J. (2020). Screening for Chinese medical staff mental health by SDS and SAS during the outbreak of Covid-19. Journal of Psychosomatic Research, 133, 110102. http://dx.doi.org/10.1016/j.jpsychores.2020.110102

Liu, C., Yang, Y., Zhang, X. M., Xu, X., Dou, Q.-L., & Zhang, W.-W. (2020). The prevalence and influencing factors for anxiety in medical workers fighting COVID-19 in China: A cross-sectional survey. *MedRxiv*, 2020.03.05.20032003. https://doi.org/10.1101/2020.03.05.20032003

Löwe, B., Wahl, I., Rose, M., Spitzer, C., Glaesmer, H., Wingenfeld, K., Schneider, A., & Brähler, E. (2010). A 4-item measure of depression and anxiety: Validation and standardization of the patient health questionnaire-4 (PHQ-4) in the general population. Journal of Affective Disorders, 122(1–2), 86–95. https://doi.org/10.1016/j.jad.2009.06.019

Montazeri, A., Vahdaninia, M., Mousavi, S. J., Asadi-Lari, M., Omidvari, S., & Tavousi, M. (2011). The 12-item medical outcomes study short form health survey version 2.0 (SF-12v2): A population-based validation study from Tehran, Iran. Health and Quality of Life Outcomes, 9(1), 12. https://doi.org/10.1186/1477-7525-9-12

Montazeri, A., Vahdaninia, M., Mousavi, S. J., & Omidvari, S. (2009). The Iranian version of 12-item short form health survey (SF-12): Factor structure, internal consistency and construct validity. BMC Public Health, 9(1), 341. https://doi.org/10.1186/1471-2458-9-341

Rohani, C., Abedi, H. A., & Langius, A. (2010). The Iranian SF-12 health survey version 2 (SF-12v2): Factorial and convergent validity, internal consistency and test-retest in a healthy sample. Iranian Rehabilitation Journal, 8(2), 4–14. http://irj.uswr.ac.ir/article-1-185-en.html

Spitzer, R., Williams, J., & Kroenke, K. Instruction manual: Instructions for patient health questionnaire (PHQ) and GAD-7 measures. Retrieved April 2, 2020, from https://phqscreeners.pfizer.edrupalgardens.com/sites/g/files/g10016261/f/201412/instructions.pdf

Spoorthy, M. S. (2020). Mental health problems faced by healthcare workers due to the COVID-19 pandemic–A review. Asian Journal of Psychiatry, 51, 102119. https://doi.org/10.1016/j.ajp.2020.102119

Ware, J. E., Kosinski, M., & Keller, S. D. (1996). A 12-Item Short-Form Health Survey: Construction of scales and preliminary tests of reliability and validity. Medical Care, 34, 220–233. http://dx.doi.org/10.1097/00005650-199603000-00003

Xiao, H., Zhang, Y., Kong, D., Li, S., & Yang, N. (2020). The effects of social support on sleep quality of medical staff treating patients with coronavirus disease 2019 (COVID-19) in January and February 2020 in China. Medical Science Monitor: International Medical Journal of Experimental and Clinical Research, 26, 923549–923549. https://doi.org/10.12659/MSM.923549

Zhang, S. X., Huang, H., & Wei, F. (2020a). Geographical distance to the epicenter of Covid-19 predicts the burnout of the working population: Ripple effect or typhoon eye effect? Psychiatry Research, 288, 112998. https://doi.org/10.1016/j.psychres.2020.112998

Zhang, S. X., Wang, Y., Rauch, A., & Wei, F. (2020b). Unprecedented disruption of lives and work: Health, distress and life satisfaction of working adults in China one month into the COVID-19 outbreak. Psychiatry Research, 288, 112958. https://doi.org/10.1016/j.psychres.2020.112958

Zhu, J., Sun, L., Zhang, L., Wang, H., Fan, A., Yang, B., Xiao, S., & Li, W. (2020). Prevalence and influencing factors of anxiety and depression symptoms in the first-line medical staff fighting against Covid-19 in Gansu. http://dx.doi.org/10.2139/ssrn.3550054

